# Laser Ablation of Periventricular Nodular Heterotopia for Medically Refractory Epilepsy

**DOI:** 10.1101/2024.02.19.24302952

**Authors:** Ryan M. McCormack, Arjun S Chandran, Samden D. Lhatoo, Sandipan Pati, Zhouxuan Li, Katherine Harris, Nuria Lacuey, Giridhar Kalamangalam, Stephen Thompson, Nitin Tandon

**Author notes:** Corresponding author: Correspondence to: Nitin Tandon, MD, FAANS, Professor, Chair *ad interim*, Department of Neurosurgery, Co-Director, Texas Institute of Restorative Neurotechnology, McGovern Medical School at UT Health, Houston, TX. 6400 Fannin Street, Suite 2800, Houston, TX, 77030.

## Abstract

**Objective:** Periventricular Nodular Heterotopia (PVNH) is the most common neuronal heterotopia, frequently resulting in pharmaco-resistant epilepsy. PVNH has a deep location which renders localization of seizure onsets and traditional surgical therapy challenging and of limited success. Here we characterize variables that predict good epilepsy outcomes following surgical intervention using SEEG-informed MRgLITT.

**Methods:** A prospectively compiled surgical epilepsy database from a single high-volume epilepsy referral center was used to identify patients who underwent SEEG evaluation for PVNH and characterize the intervention on outcomes.

**Results:** Thirty-nine patients underwent SEEG-informed MRgLITT. Associated imaging abnormalities— mesial temporal sclerosis (MTS) or polymicrogyria (PMG) were treated based on SEEG.

SEEG-guided MRgLITT of the seizure onset zone (SoZ) in PVNH and associated epileptic tissue was carried out. PVNH and PMG were densely sampled—mean 16.5(SD=2)/209.4(SD=36.9) SEEG probes/recording contacts. A single trajectory was used in 18, two in 13, and three or more in eight patients. Volumetric analyses revealed a high percentage of PVNH SoZ ablation (96.6%, SD=5.3%) in unilateral and bilateral (92.9%, SD=7.2%) cases. Mean follow-up duration was 31.4 months (SD=20.9). Seizure freedom was excellent overall: unilateral PVNH without other imaging abnormalities—80%; PVNH with MTS or PMG—63%; Bilateral PVNH—50%. SoZ ablation percentage significantly impacted surgical outcomes (*p*<0.001).

**Interpretation:** PVNH plays a central role in seizure genesis. MRgLITT represents a transformative technological advance in PVNH-associated epilepsy with seizure control outcomes consistent with those seen in focal lesional epilepsies. In localized unilateral cases and otherwise normal imaging, performing PVNH ablation without invasive recordings may be reasonable.

## Introduction

Periventricular Nodular Heterotopia (PVNH) is a neuronal migration disorder characterized by ectopic neuronal rests in subependymal locations due to aberrant migration. It’s expression varies from a single focal periventricular nodule to large diffuse bilateral nodules,^1^ with normal, polymicrogyric, or pachygyric overlying cortex.^2-6^ Most sporadic cases are not associated with germ-line mutations of the kind seen in X-linked dominant Filamin 1 mutation in familial PVNH.^7^ Many forms of PVNH result in drug-resistant seizures that are thought to variably originate in PVNH,^1,8^ overlying cortex and/or mesial temporal structures.^2,4,7,9-11^ Standard microsurgical approaches are impractical in PVNH owing to deep location, often large extent, and location close to critical subcortical structures which increase risks of collateral damage. The prevailing standard for PVNH-associated intractable epilepsy is to record from nodules, overlying cortex, and other components of the putative seizure network using SEEG, followed by thermo-coagulation, radiosurgery, or neuromodulation.^12-16^ Outcomes from these approaches have been inferior to other lesional epilepsies (e.g. focal cortical dysplasia, cavernous malformations), where high cure rates are readily achieved.

MR-guided laser interstitial thermal therapy (MRgLITT) enables highly precise targeting of PVNH.^17^ Initial reports of MRgLITT for PVNH have been limited by small numbers,^18,19^ rendering it difficult to categorize outcomes and isolate predictive factors.^20^ Higher-density intracranial recordings may enable more precise targeting of the PVNH seizure network.^7,10,21^ We asked the following questions: Is LlTT safe and effective in this disorder. Does it work even if the disease is bilateral and/or associated with other developmental anomalies like PMG? Can we treat PVNH as a lesional epilepsy without first proving that nodules are the site of seizure genesis? In these cases, do outcomes correlate with extent of ablation? To address these questions, we studied thirty-nine patients who underwent SEEG (with multiple PVNH recodings sites) and subsequent MRgLITT (with multiple fiber trajectories to enable comprehensive ablation). This comprises the largest MRgLITT surgical experience to date for PVNH-associated epilepsy.

## Materials and Methods

### Study Participants

A prospectively maintained database of epilepsy patients was used to identify all patients with PVNH-associated epilepsy that underwent surgical intervention. We then focused on patients that underwent MRgLITT of the PVNH nodule(s) causing epilepsy. All patients were diagnosed with medically resistant epilepsy, defined by resistance to three or more anti-seizure medications (ASMs). All study procedures were performed after approval from the local committee for the protection of human subjects.

### Epilepsy Evaluation

The pre-surgical evaluation included a neurologic history, physical examination, inpatient scalp video electroencephalogram (EEG), epilepsy protocol 3-Tesla MR imaging, and standardized neuropsychological testing. 2-[18F] fluoro-2-deoxy-D-glucose positron emission tomography (FDG-PET), intracarotid amytal test, magnetoencephalography (MEG), and functional MRI testing were performed as deemed necessary. Intra-cranial recordings were performed using previously published SEEG techniques.^22,23^ All data were reviewed and surgical decisions made at a multi-disciplinary epilepsy management conference. Interventions before SEEG were considered unconnected to the current analysis, that is focused on characteristics and interventions after SEEG evaluation. (Supplementary Table 3).

### Surgical Procedure

SEEG recordings were performed using 0.8-mm diameter electrodes (PMT Corp, Chanhassen MN) implanted using a ROSA robot (Zimmer Biomet, Warsaw IN) guided by a computed tomographic angiogram and high-resolution 3D-contrasted T1-weighted MRI scans.^23,24^ MRgLITT was performed using a Medtronic Visualase system (Medtronic, Minneapolis MN). Stereotactic planning (Medtronic Stealth S8 workstation) was used to optimize laser trajectories for maximal transversal of PVNH as detailed previously.^18,23,25,26^ Seizure onset electrodes were imported as separate image sets to enable definitive targeting within the ablation zone. The risk of injury to visual radiations was minimized using either tractography of visual radiations or anatomical landmarks relative to the ventricular system. If other cortical malformations (PMG, MTS) were implicated in seizure genesis, they were also ablated, either at the same or subsequent session or with microsurgical resection. Ablation was optimized using real-time thermal imaging in a 3T MRI scanner.^25^ Post-ablation MRI imaging, scalp EEG, formal visual field testing, and neuropsychological assessment were obtained at 6 months post-op. Post-surgical seizure outcomes were categorized using ILAE and Engel scales.

### Imaging analysis

Pre-ablation high-resolution T1 and T2 imaging and post-ablation high-resolution T1 with contrast imaging sequences were co-registered. Seizure onset electrodes from SEEG recordings were overlayed to identify the lesional seizure onset zone (SoZ). Pre- and postsurgical 3D volumetric estimates were performed after manual contouring of total lesion (PVNH or PMG), lesional SoZ, and ablation zone in iPlan software (Brainlab). Statistical analyses were performed using SPSS 28.0 (IBM). Fisher’s exact test was for univariate and multivariate analysis.

## Results

### Patient Characteristics

Seven hundred and ninety-four patients underwent neurosurgical treatment for epilepsy at our center between February 2006 to February 2022 by the senior author. Of these, fifty-two underwent intracranial evaluations for PVNH. Ten underwent subdural electrode implantations followed by microsurgical resection of (temporal or temporo-occipital) PVNH, while forty-two underwent an SEEG evaluation. Three patients undergoing SEEG did not undergo MRgLITT but instead underwent neuromodulation (two) or a topectomy (one). The thirty-nine patients with PVNH (Table 1, Supplementary Table 1) that underwent SEEG-informed MRgLITT (6/2012 – 2/2022) form the basis of this analysis. Average age at surgery was 30 years (range: 16-51 years), average age at epilepsy onset was 14.9 years (range: 1-40 years), and average time between seizure onset and intervention was 15.1 years (range: 1-41 years). The cohort comprised 28 females (71.8%) (Supplementary Table 1). The average follow-up after definitive intervention was 31.4 months (range: 8 - 85 months). 37/39 patients were followed for at least one year (Supplementary Table 2). Fourteen patients had another epilepsy surgical procedure before PVNH ablation, with six having the procedure before SEEG (five – ATL, four - mesial temporal MRgLITT, two - prior cortical topectomy, three - VNS placement) (Supplementary Table 3). Of these, thirteen (92.9%) had poor epilepsy surgical outcomes before PVNH-LITT ablation. The patient with a good initial outcome following surgery but before PVNH-LITT (Patient 1) had very short-term f/u as he underwent PVNH-LITT two months after ATL. Two patients underwent additional epilepsy surgery after PVNH-LITT (ATL, Topectomy), both of which were planned preoperatively based on SEEG and not by breakthrough seizures (Supplementary Table 3).

**Table 1:** Patients with unilateral and bilateral PVNH – Imaging and SEEG characteristics.

|  | Unilateral | Bilateral | Total |
| --- | --- | --- | --- |
| Imaging (PVNH) | 15 | 24 | 39 |
| • Localized |  |  |  |
| • Frontal | 2 | - | 2 |
| • Occipital | 2 | - | 2 |
| • Temporal | 2 | - | 2 |
| • Temporal, Occipital | 5 | 6 | 11 |
| • Temporal, Occipital, Parietal | 2 | 3 | 5 |
| • Frontal, Parietal, Occipital | 1 | - | 1 |
| • Temporal, Parietal | 1 | 1 | 2 |
| • Frontal, Parietal | - | 3 | 3 |
| • Frontal, Temporal, Parietal | - | 1 | 1 |
| • Parietal, Occipital | - | 1 | 1 |
| • Diffuse | - | 9 | 9 |
| Cortical Abnormality |  |  |  |
| • PMG | 3 | 4 | 7 |
| • MTS | 3 | 6 | 9 |
| • PMG + MTS | - | 4 | 4 |
| • None | 10 | 10 | 19 |
| Seizure Onset Zone (PVNH)* | 28 | 11 | 39 |
| Cortical Abnormality in SoZ* |  |  |  |
| • PMG | 2 | 3 | 5 |
| • MTS | 7 | 0 | 7 |
| • PMG + MTS | 2 | 0 | 2 |
| • None | 15 | 8 | 21 |
| Primary SEEG finding (SoZ)*: | -- | -- | -- |
| • Low voltage fast | 20 | 6 | 26 |
| • Multiple<sup>†</sup> | 6 | 5 | 11 |
| • Rhythmic alpha | 1 | - | 1 |
| • Semi-rhythmic beta | 1 | - | 1 |

\* Only including patients that underwent PVNH LITT
<sup>†</sup> Multiple include low voltage fast

### Non-invasive data

Fifteen patients had unilateral (seven right, eight left), fifteen had bilateral localized, and nine had diffuse bilateral multi-lobar PVNH (Table 1, Supplementary Table 1, Supplementary Fig. 1). There was a clear preponderance of nodules in paratrigonal regions of lateral ventricles (51.3% - 20/39). Nineteen (48.7%) had abnormal MRI findings besides PVNH: polymicrogyria (seven), mesial temporal abnormalities (eight), or both (four) (Table 1, Supplementary Table 1, Supplementary Fig. 1). Four predominant seizure types were identified by SEEG (Supplementary Table 1). Focal to bilateral tonic-clonic (FTBTC) were the commonest (n=22) seizures; focal impaired awareness seizures (FIAS) were second most common (n=ten). The two other seizure types were FIAS with rare FTBTC (n=five) and only visual aura (n=two).

### Invasive Monitoring

Of the fifteen SEEG patients with unilateral focal PVNH, three underwent bilateral SEEG due to discordance on scalp EEG or other imaging findings (Supplementary Table 1, 2), and the others underwent a unilateral implantation based on imaging and semiology. Of the twenty-four patients with bilateral PVNH (localized or diffuse), six had unilateral onsets on scalp EEG (Supplementary Table 1). However, all underwent bilateral invasive monitoring, given bilateral imaging abnormalities. Across all patients (n= 39), the mean number of SEEG probes implanted was 16.6 (SD = 1.9), with an average of 209.4 (SD = 36.9) recording contacts. In cases of unilateral SEEG (n=12), the average number of probes was 14.1 (SD = 1.7), with an average of 160.5 (SD = 11) recording contacts. In bilateral implantations (n = 27) the mean number of probes was 17.6 (SD = 1.1), with an average of 235.3 (SD = 12.2) contacts. Representative cases are shown in Figures 2A-F and 2G-N. The predominant ictal onset rhythm from nodules was low voltage fast (twenty-six patients), rhythmic alpha (one patient), and semi-rhythmic beta (one patient) (Table 1). Multiple types of ictal onset patterns, including low-voltage fast activity, were identified in the other eleven patients (Table 1). Previous studies have highlighted a predominant pattern of low-voltage fast in PVNH; however, definitive statements about this pattern have been limited by low patient numbers.^16,19^ We found that low-voltage fast seizure onset was either the lone pattern or part of multiple patterns in 37 patients (26 only low voltage fast, 11 multiple), suggesting this may be a biomarker of epileptogenic PVNH.

**Figure 1:**
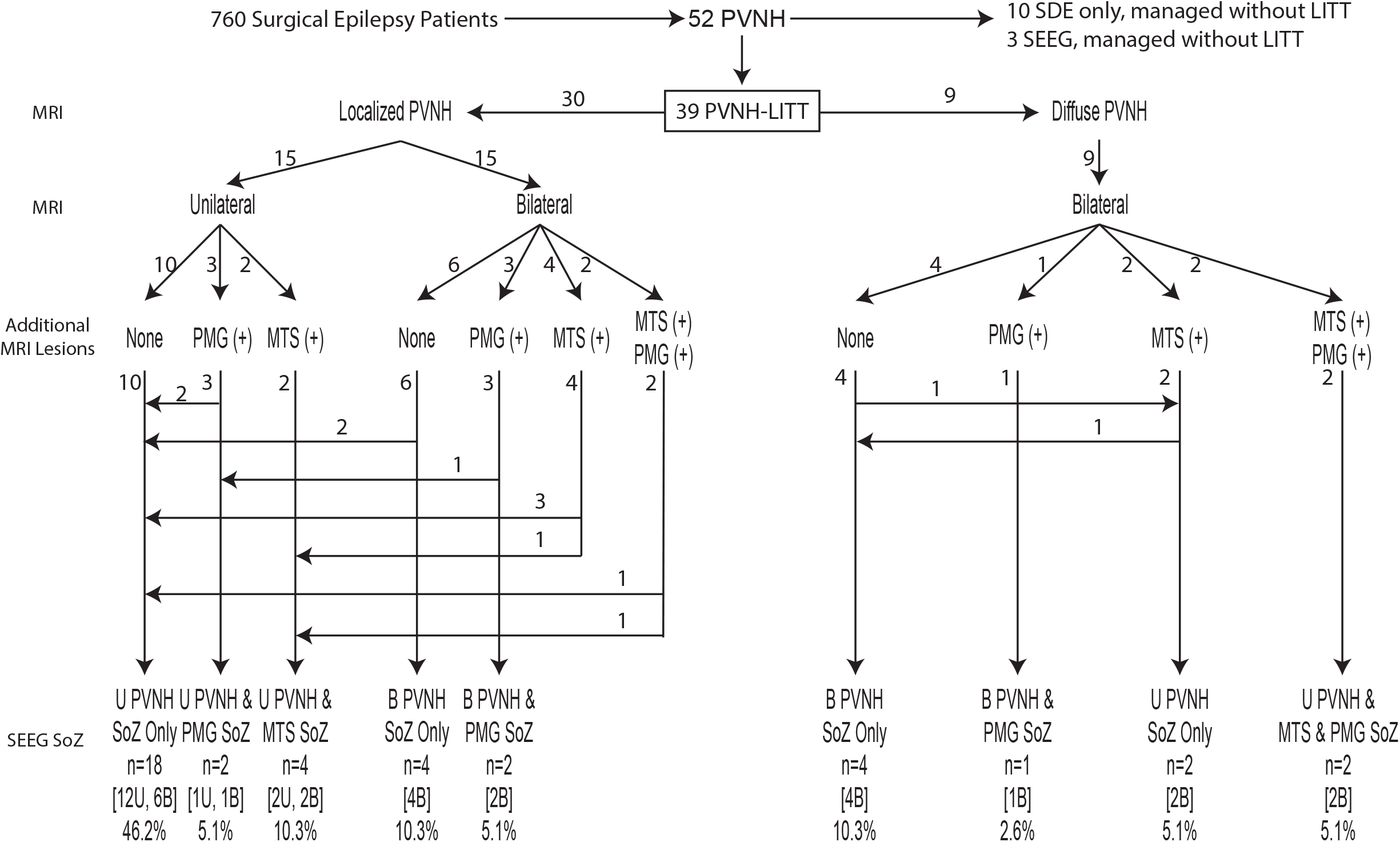
Characterization of patients based on imaging. Patients were first grouped based on local or diffuse MRI findings, plus the presence or absence of other imaging anomalies on MRI, and then coupled with seizure onset zone localization using SEEG. Brackets indicate PVNH localization from MRI, U = Unilateral, B = Bilateral.

**Figure 2:**
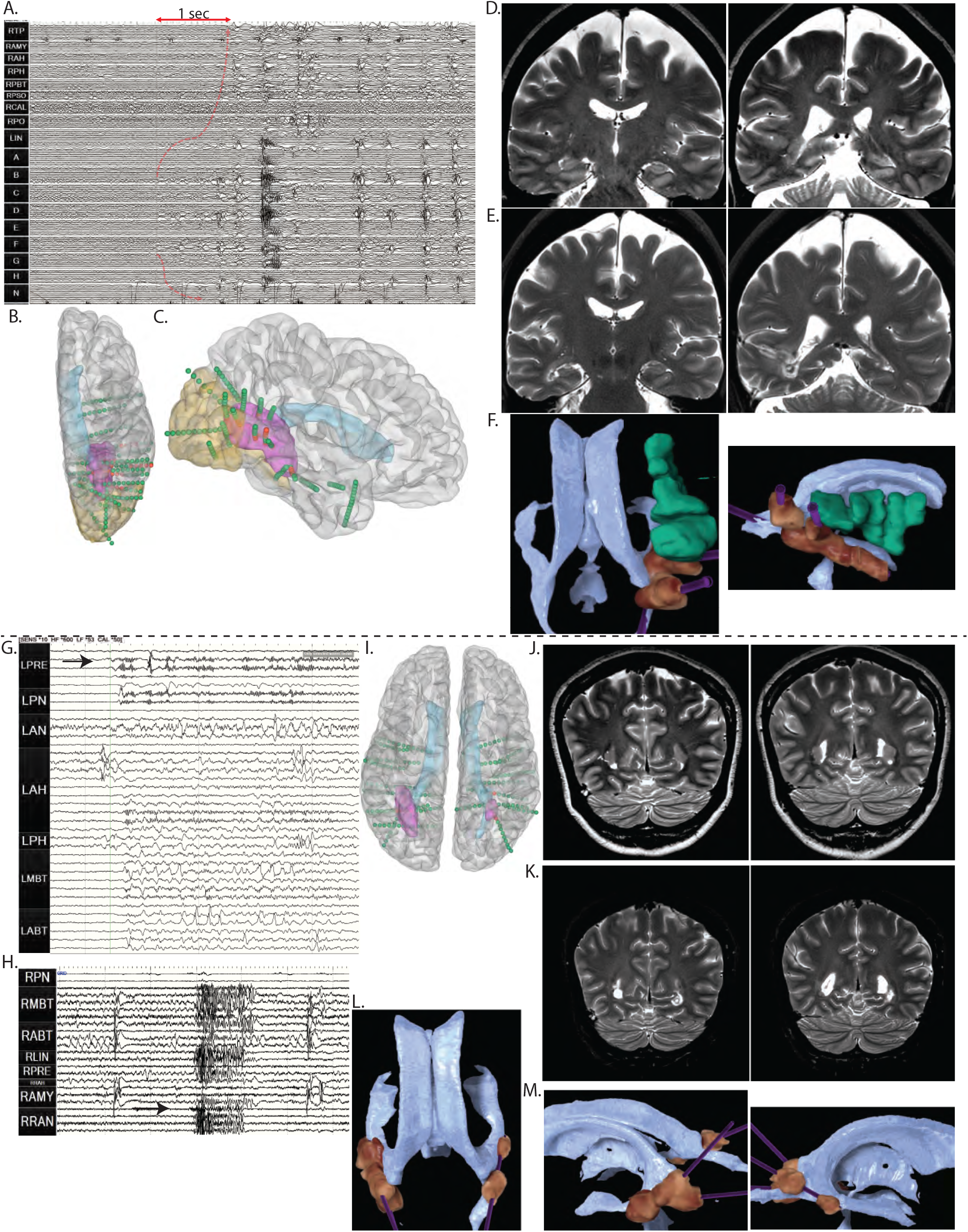
Exemplars of patients with unilateral or bilateral PVNH: A patient with unilateral localized PVNH and PMG (A-F) and bilateral localized PVNH (G-M). A: SEEG recordings demonstrating PVNH onset in PVNH nodule electrodes followed by cortical spread. B, C: Electrodes (green) co-localized with PVNH (pink), PMG (orange), and ventricle (blue). (D, E) Pre- (D) and 6 months post-ablation (E) of patient SoZ PVNH and PMG as informed by SEEG. (F) Volumetric visualization of ablated PVNH and PMG (brown). Ventricle (blue), MRgLITT fiber (purple), and PMG (Green). G, H: SEEG recordings reveal a PVNH onset followed by cortical spread. I: Electrodes (green) relative to PVNH (purple) and ventricle (blue). J, K: Pre- and 6-month post-ablation images of PVNH as informed by SEEG. L, M: Volumetric visualization of ablated PVNH (brown), ventricle (blue), MRgLITT fiber (purple).

Seizure onsets were either localized to PVNH (*n*=21) or involved a combination of nodules and cortex (*n*=18). In cases of combined nodular and cortical onsets, 11 had additional MRI abnormalities (i.e. PMG or MTS) where onsets were also localized, whereas seven patients had no imaging lesions besides PVNH. The number of recording contacts in PVNH correlated with the ability to discern if the SoZ comprised PVNH only or PVNH plus overlying cortex (*P*=0.003). In the 32 cases where the PVNH SoZ was localized to only the PVNH nodule, there were an average of 19.8 (SD 8.6) recording contacts in PVNH. In comparison, there were significantly fewer contacts in the seven cases where the SoZ included cortex and nodule with an average of 8.7 (SD 5.4) PVNH recording contacts (*P*=0.003). For patients with PVNH, no other MRI visible lesion, and additional non-PVNH cortical onsets, only 43% (3/7) had good surgical seizure outcomes with ablation of PVNH. The three good responders had a significantly higher PVNH SEEG sampling than the four poor responders (14.3 vs 4.5 PVNH contacts, *P*<0.005). Subdural grids (SDE) were used as an adjunct to SEEG (separate operation after SEEG explantation) for decision-making in five early patients with PVNH (four left, one right SDE) who had prominent cortical involvement in addition to PVNH onset. 80% (4/5) had durable seizure freedom, and all had good seizure outcomes.

Of the three patients who underwent SEEG but were not candidates for PVNH MRgLITT, one had occipital FCD + HS, one had HS, and one had schizencephaly. One had only cortical onsets, while two had multifocal cortical onsets with interictal activity in the PVNH (Supplementary Table 1). PVNH was thus not directly targeted with MRgLITT in these three patients due to minimal PVNH involvement in seizure genesis (Patients 14 and 18) or multifocal bilateral onsets prompting neuromodulation (RNS) (Patient 6).

### Interventions and Seizure Outcomes

We assumed that PVNH adjacent to the specific SEEG electrode showing seizure onset was interwoven with the seizure onsets and so all thirty-nine patients underwent MRgLITT of the maximal amount of PVNH proximate to the SoZ. 18 underwent ablation via a single probe. In 13 patients, two probes were used. Three underwent ablation using three probes, and in five patients, four probes were used (Supplementary Table 2). Seventeen patients underwent left-sided, eleven underwent right-sided, and another eleven had bilateral ablations over one (3 patients) or two sessions (8 patients) (Figures 2E-G and K-M). Only two (5.1%) patients underwent another non-LITT surgical intervention (ATL or a topectomy) following MRgLITT of PVNH (Supplementary Table 3).

The percentage of the SoZ within PVNH that was ablated significantly correlated (*P*<0.05) with good surgical seizure outcomes, both in unilateral and bilateral PVNH (Figure 3D). Volumetric analyses of residual PVNH following MRgLITT revealed that in unilateral cases (*n*=28), SoZ ablation averaged 95.6% (95% CI 93.4-97.8), and in bilateral cases (*n*=11), it averaged 92.9% (95% CI 88.6-97.2). There was a significant difference (*P*<0.05) in the average SoZ ablation in cases with good vs. poor outcomes in both unilateral (96.8% vs 80.8%) and bilateral (95% vs 82.3%) cases (Figure 2D). In the six cases where PMG was involved in the SoZ, the proportion of SoZ ablated averaged 91.4% (95% CI 87.1 – 95.8). (Figure 3, Supplementary Table 2, Supplementary Table 4, 5). All patients undergoing PMG ablation (four concurrent with PVNH MRgLITT, one before PVNH MRgLITT) had good surgical seizure outcomes (Figure 1, Table 1, Supplementary Table 2). No patients had SAH MRgLITT at the same time as PVNH MRgLITT in this series.

**Figure 3:**
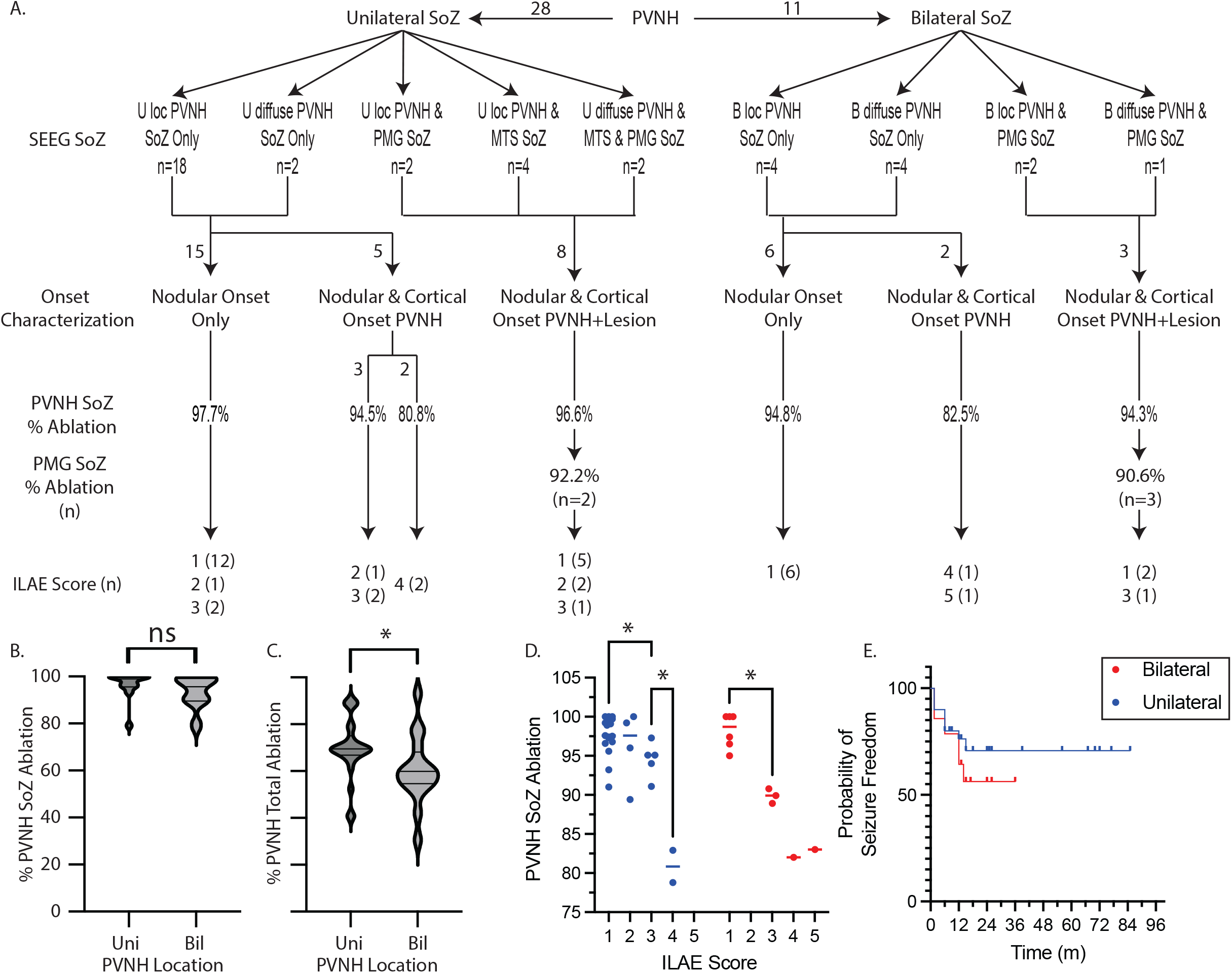
Impact of extent of the seizure onset zone and multiple pathologies on seizure outcomes. A: SoZ revealed by SEEG in either PVNH only or in PVNH plus cortical onset with average SoZ percentage ablation of PVNH or PMG for each group related to ILAE score at the most recent follow-up. B: Percentage PVNH SoZ ablation for bilateral and unilateral PVNH onsets. C: Percentage of total PVNH on imaging ablation for bilateral and unilateral onsets. D: Scatter plot relating the percentage PVNH SOZ ablation to ILAE score at the last follow-up. E: Survival analysis of seizure freedom following PVNH ablation. * p<0.05, Red – bilateral PVNH onsets, blue – unilateral PVNH onsets.

As might be expected, there was a significantly greater (*P*<0.05) (76%) proportion of PVNH ablated in unilateral compared to bilateral cases (66.2%); however, there was not a significant difference in good surgical outcomes following total PVNH ablation in either unilateral (76.4% good vs 70.6% poor) or bilateral cases (67.3% good vs 59.9% poor) (Supplementary Table 4, 5). PVNH MRgLITT led to a significant reduction in ASM usage (p<0.05). ASMs were reduced in 37 patients and eight patients (20.5%) were weaned off ASMs completely. 90.5% (19/21) of patients with PVNH-only onsets were seizure-free and all had good surgical outcomes following MRgLITT. 81.8% (9/11) of patients with cortical and PVNH onsets with cortical abnormalities on imaging were seizure-free and all had good surgical outcomes following MRgLITT (Figure 3, Supplemental Table 3). Seizure outcomes were durable following MRgLITT with 89.2% (33/37), having good seizure outcomes (Engel 2 /ILAE 3 or better) at 12 months. Correspondingly, seizure-free rates (ILAE 2 or better) were also high – 73.1% in unilateral (19/26) and 72.7% in bilateral cases (8/11) at 12 months (Figure 3E).

Seven patients demonstrated both nodular and cortical onsets but had no visible cortical abnormalities by MRI (Figure 3). In these seven, only PVNH was ablated with no additional cortical ablation. Four of these seven patients had poor surgical outcomes (Engel III or IV; ILAE >3). These four had bilateral PVNH (two localized, two diffuse). PVNH SoZ ablation percentage in good responders (*n*=35) was also significantly smaller (*P*<0.0001) as compared to poor responders (*n*=four) (Figure 3D). In aggregate, this suggests that in diffuse or widespread disease, SEEG is helpful for accurately identifying the SoZ.

Six patients (15.4%) had prior epilepsy surgery before SEEG and eight (20.5%) underwent epilepsy surgery following SEEG besides PVNH MRgLITT (Supplementary Table 3), based on staged plans developed after integration of the intracranial EEG data. Seven out of eight patients had poor surgical seizure outcomes until PVNH ablation, reiterating that PVNH is the major driver of the epileptogenic network (Supplementary Table 3) and should be targeted aggressively with recordings and ablation.

### Complications

Three patients suffered cortical hemorrhages, one each following either SEEG implantation, SEEG explantation, or MRgLITT explantation. The SEEG implantation hemorrhage was discovered incidentally on routine post-operative imaging and the two explantation complications experienced post-operative seizures prompting imaging. Each complication was managed non-operatively with one extra day of inpatient observation and none required additional intervention. Five patients developed new visual field deficits following PVNH MRgLITT. Two patients developed an octanopsia,^27^ and three developed a quadranopsia. None of these visual deficits were clinically relevant and did not preclude driving in seizure-free patients.

## Discussion

PVNH has hitherto been a highly intractable and challenging epilepsy syndrome. The advent of MRgLITT opens an entirely new avenue to managing these patients. Our prior report of four PVNH-associated epilepsy cases and others have described six patients who underwent MRgLITT, have been the only reports on the subject so far.^18,20^ We extend our cohort to thirty-nine patients, with longer follow-up (mean 32.9 months, 37 with greater than one-year of follow-up), and report good outcomes (ILAE 3 or better) in 89%. Thus, MRgLITT represents a transformative technological advance in the management of PVNH.

### Intracranial recordings and implication of PVNH in epilepsy network

The epilepsy network in PVNH has been hypothesized to be widely distributed and complex.^7,10,14,19^ The relative paucity of publications relating to surgical management of these malformations has left questions about the epileptogenic role of the nearby cortex unanswered. Our series illustrates the central role of the PVNH in seizure genesis. The impact of the percentage of ablation on outcome points to the criticality of eliminating this abnormality in enabling seizure freedom.^19^ Further, even in cases with simultaneous nodular and cortical onset, good results are achieved simply by nodule ablation, indicating a pivotal role of the PVNH in seizure generation when no other lesions are present.^7,10,14,19^ Furthermore, when imaging abnormalities are restricted to PVNH, good epilepsy surgical outcomes appear mostly related to the extent of PVNH ablation. Intracranial recordings in diffuse or bilateral cases are warranted to delineate the epileptic PVNH network for targeted ablation. A high density of recording electrodes enabling thorough PVNH coverage may explain reveal predominantly nodular or combined cortical-nodular onsets, a divergence from other published reports.^7,20^ Tassi *et al*.^7^ reported seizures from the PVNH nodule and overlying cortex but never only from nodules; however, only 12.4 probes were used per patient (compared to our 16.5).^7^ Durica *et al*.^20^ also identified 4/6 patients having seizure onset from the PNVH nodule with an SEEG scheme with good nodule coverage - an average of 16 electrodes and 192 recording contacts per patient.^20^ These findings suggest that dense SEEG coverage may reveal focal nodular onsets more reliably and enable targeted ablation in patients with widespread or diffuse PVNH.

While our approach has been to perform SEEG before MRgLITT for PVNH, this cohort supports a hypothesis that select patients could benefit from ablation *without* the need for invasive intracranial recordings. This opinion is supported by the concordance of seizure onset zones with PVNH, the high percentage of ablation of aberrant tissue in localized PVNH (>90%), and safety and efficacy of MRgLITT. We propose an algorithm for managing patients with unilateral localized PVNH and no cortical abnormalities using MRgLITT without prior SEEG as a first option (Figure 4). Figure 4B demonstrates that there are no significant differences detected between the ablation of unilateral localized PVNH based on MRI imaging or the SEEG-guided SoZ. In contrast, SEEG evaluations are necessary to further evaluate the seizure network in cases with extensive PVNH or other cortical abnormalities on imaging. Given the intensive healthcare resource utilization and potential for morbidity with SEEG, this approach carries major implications for epilepsy surgery access, particularly in resource-deprived populations.^28^

**Figure 4:**
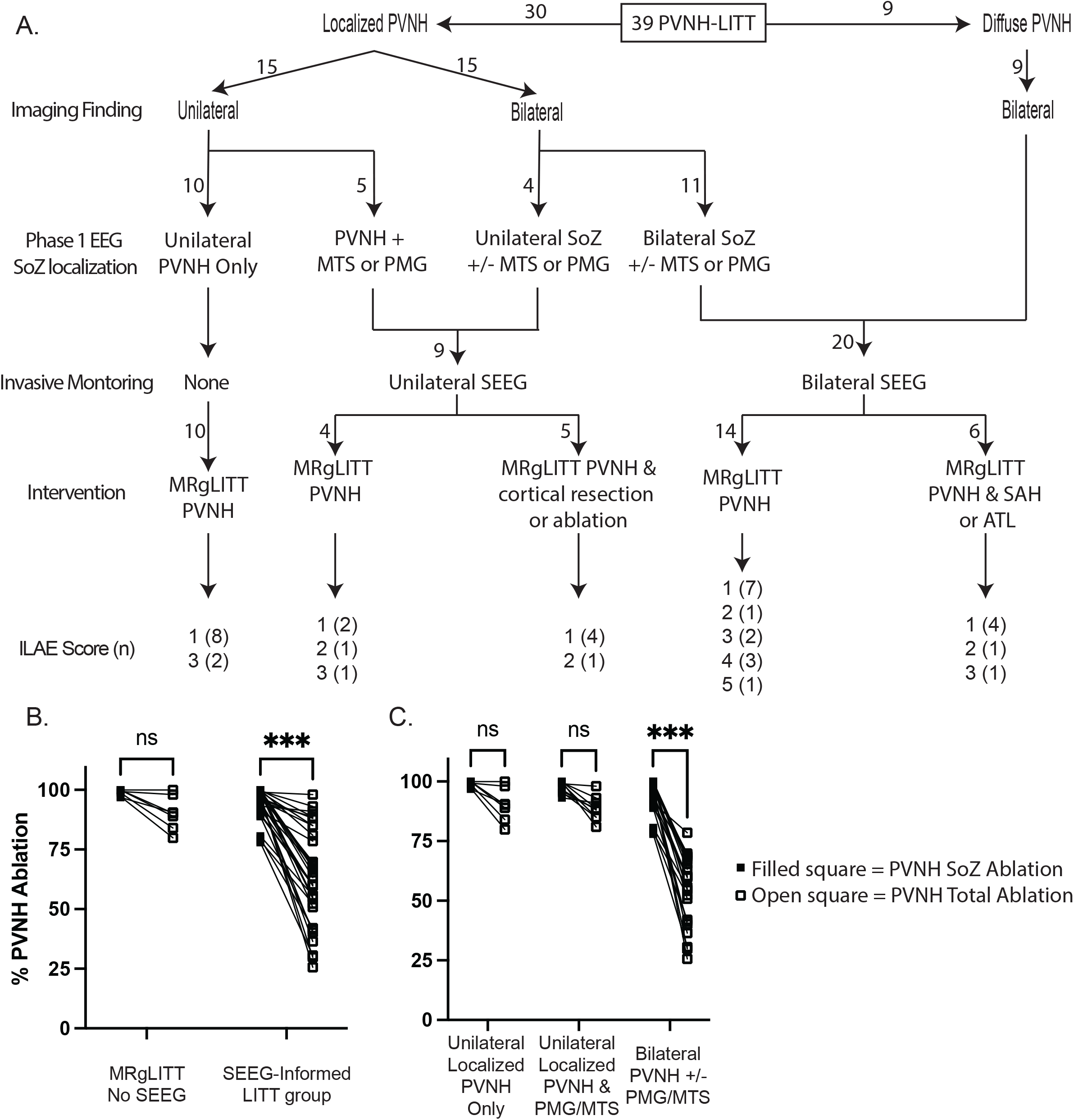
SEEG may be superfluous in unilateral PVNH without other imaging abnormalities. A: A proposed decision tree for evaluation and intervention in patients with PVNH-associated epilepsy. Patient numbers are based on a retrospective analysis of the patient cohort in this study. B, C: Unilateral localized PVNH does not significantly impact ablation percentage. Pairwise graphed comparison between SEEG-informed seizure onset zone ablation (closed circle) and total PVNH volume ablated (open circle) for (B) proposed “no SEEG” population compared to the population where SEEG is recommended for further evaluation and (C) Unilateral localized PVNH with or without cortical abnormality compared to bilateral PVNH ablation. Ns – not significant *** p < 0.001.

## Conclusion and Limitations

MRgLITT for PVNH-associated epilepsy is a safe, feasible, and highly efficacious intervention with good surgical outcomes. In focal localized PVNH with no other lesions and generally concordant EEG, laser ablation of PVNH may be considered a first-line treatment option. When SEEG is indicated (bilateral, widespread PVNH) high-density implantation helps with localization. The validation of a MRgLITT-only approach, sans SEEG in focal isolated PVNH, merits validation in a prospective cohort.

## Data Availability

All data produced in the present work are contained in the manuscript

## Abbreviations

PVNH: Periventricular Nodular Heterotopia
SEEG: Stereoelectroencephalography
MRgLITT: Magnetic Resonance-Guided Laser Interstitial Thermal Therapy
MTS: Mesial Temporal Sclerosis
PMG: Polymicrogyria
SoZ: Seizure Onset Zone
ASM: Antiseizure Medication
FDG-PET: 2-[18F] fluoro-2-deoxy-D-glucose positron emission tomography
MEG: magnetoencephalography
ATL: Anterior Temporal Lobectomy
VNS: Vagal Nerve Stimulator
FTBTC: Focal to bilateral tonic-clonic
FIAS: focal impaired awareness seizures
SDE: Subdural Grid Electrode
SAH: Selective Amygdalohippocampectomy
FCD: Focal Cortical Dysplasia
HS: Hippocampal Sclerosis
RNS: Responsive Neurostimulation
VNS: Vagal Nerve Stimulator

## Figure Legends

Supplemental Figure 1: Imaging distribution of PVNH patients. Distribution and characterization of patients based on strictly imaging findings from cranial MRI. Patients are distributed based on the type of PVNH (Localized or Diffuse) and location. Patients are subsequently distributed based on the presence of mesial temporal sclerosis (MTS) or polymicrogyria (PMG).

## Author Contributions

SDL, SP, KH, NL, GK, ST, NT contributed to conception and design of the study. RM, SL, SP, ZL, KH, NL, GK, ST, NT contributed to acquisition and analysis of the data. RM, SL, SP, ST, NT contributed to drafting the text or preparing the figures.

## References

1. Broix L, Jagline H, Ivanova E, et al. Mutations in the HECT domain of NEDD4L lead to AKT-mTOR pathway deregulation and cause periventricular nodular heterotopia. Nat Genet. Nov 2016;48(11):1349-1358. doi:10.1038/ng.3676

2. Hannan AJ, Servotte S, Katsnelson A, et al. Characterization of nodular neuronal heterotopia in children. Brain. Feb 1999;122 (Pt 2):219–38. doi:10.1093/brain/122.2.219

3. Kakita A, Hayashi S, Moro F, et al. Bilateral periventricular nodular heterotopia due to filamin 1 gene mutation: widespread glomeruloid microvascular anomaly and dysplastic cytoarchitecture in the cerebral cortex. Acta Neuropathol. Dec 2002;104(6):649–57. doi:10.1007/s00401-002-0594-9

4. Guerrini R, Parrini E. Neuronal migration disorders. Neurobiol Dis. May 2010;38(2):154–66. doi:10.1016/j.nbd.2009.02.008

5. Wieck G, Leventer RJ, Squier WM, et al. Periventricular nodular heterotopia with overlying polymicrogyria. Brain. Dec 2005;128(Pt 12):2811–21. doi:10.1093/brain/awh658

6. Oegema R, Maat-Kievit A, Lequin MH, et al. Asymmetric polymicrogyria and periventricular nodular heterotopia due to mutation in ARX. Am J Med Genet A. Jun 2012;158A(6):1472–6. doi:10.1002/ajmg.a.35365

7. Tassi L, Colombo N, Cossu M, et al. Electroclinical, MRI and neuropathological study of 10 patients with nodular heterotopia, with surgical outcomes. Brain. Feb 2005;128(Pt 2):321–37. doi:10.1093/brain/awh357

8. Fallil Z, Pardoe H, Bachman R, et al. Phenotypic and imaging features of FLNA-negative patients with bilateral periventricular nodular heterotopia and epilepsy. Epilepsy Behav. Oct 2015;51:321–7. doi:10.1016/j.yebeh.2015.07.041

9. Francione S, Kahane P, Tassi L, et al. Stereo-EEG of interictal and ictal electrical activity of a histologically proved heterotopic gray matter associated with partial epilepsy. Electroencephalogr Clin Neurophysiol. Apr 1994;90(4):284–90. doi:10.1016/0013-4694(94)90146-5

10. Battaglia G, Chiapparini L, Franceschetti S, et al. Periventricular nodular heterotopia: classification, epileptic history, and genesis of epileptic discharges. Epilepsia. Jan 2006;47(1):86–97. doi:10.1111/j.1528-1167.2006.00374.x

11. Kothare SV, VanLandingham K, Armon C, Luther JS, Friedman A, Radtke RA. Seizure onset from periventricular nodular heterotopias: depth-electrode study. Neurology. Dec 1998;51(6):1723–7. doi:10.1212/wnl.51.6.1723

12. Pizzo F, Roehri N, Catenoix H, et al. Epileptogenic networks in nodular heterotopia: A stereoelectroencephalography study. Epilepsia. Dec 2017;58(12):2112–2123. doi:10.1111/epi.13919

13. Cossu M, Mirandola L, Tassi L. RF-ablation in periventricular heterotopia-related epilepsy. Epilepsy Res. May 2018;142:121–125. doi:10.1016/j.eplepsyres.2017.07.001

14. Mirandola L, Mai RF, Francione S, et al. Stereo-EEG: Diagnostic and therapeutic tool for periventricular nodular heterotopia epilepsies. Epilepsia. Nov 2017;58(11):1962–1971. doi:10.1111/epi.13895

15. Wu C, Sperling MR, Falowski SM, et al. Radiosurgery for the treatment of dominant hemisphere periventricular heterotopia and intractable epilepsy in a series of three patients. Epilepsy Behav Case Rep. 2013;1:1–6. doi:10.1016/j.ebcr.2012.10.004

16. Nune G, Arcot Desai S, Razavi B, et al. Treatment of drug-resistant epilepsy in patients with periventricular nodular heterotopia using RNS(R) System: Efficacy and description of chronic electrophysiological recordings. Clin Neurophysiol. Aug 2019;130(8):1196–1207. doi:10.1016/j.clinph.2019.04.706

17. Youngerman BE, Save AV, McKhann GM. Magnetic Resonance Imaging-Guided Laser Interstitial Thermal Therapy for Epilepsy: Systematic Review of Technique, Indications, and Outcomes. Neurosurgery. Apr 1 2020;86(4):E366–E382. doi:10.1093/neuros/nyz556

18. Esquenazi Y, Kalamangalam GP, Slater JD, et al. Stereotactic laser ablation of epileptogenic periventricular nodular heterotopia. Epilepsy Res. Mar 2014;108(3):547–54. doi:10.1016/j.eplepsyres.2014.01.009

19. Thompson SA, Kalamangalam GP, Tandon N. Intracranial evaluation and laser ablation for epilepsy with periventricular nodular heterotopia. Seizure. Oct 2016;41:211–6. doi:10.1016/j.seizure.2016.06.019

20. Durica SR, Caruso JP, Podkorytova I, et al. Stereo-EEG Evaluation and Surgical Treatment in Patients With Drug-Resistant Focal Epilepsy Associated With Nodular Heterotopia. J Clin Neurophysiol. Apr 7 2021;doi:10.1097/WNP.0000000000000850

21. Aghakhani Y, Kinay D, Gotman J, et al. The role of periventricular nodular heterotopia in epileptogenesis. Brain. Mar 2005;128(Pt 3):641–51. doi:10.1093/brain/awh388

22. Rollo PS, Rollo MJ, Zhu P, Woolnough O, Tandon N. Oblique trajectory angles in robotic stereo-electroencephalography. J Neurosurg. Aug 14 2020:1–10. doi:10.3171/2020.5.JNS20975

23. Tandon N, Tong BA, Friedman ER, et al. Analysis of Morbidity and Outcomes Associated With Use of Subdural Grids vs Stereoelectroencephalography in Patients With Intractable Epilepsy. JAMA Neurol. Jun 1 2019;76(6):672–681. doi:10.1001/jamaneurol.2019.0098

24. Gonzalez-Martinez J, Bulacio J, Thompson S, et al. Technique, Results, and Complications Related to Robot-Assisted Stereoelectroencephalography. Neurosurgery. Feb 2016;78(2):169–80. doi:10.1227/NEU.0000000000001034

25. Donos C, Breier J, Friedman E, et al. Laser ablation for mesial temporal lobe epilepsy: Surgical and cognitive outcomes with and without mesial temporal sclerosis. Epilepsia. Jul 2018;59(7):1421–1432. doi:10.1111/epi.14443

26. Wicks RT, Jermakowicz WJ, Jagid JR, et al. Laser Interstitial Thermal Therapy for Mesial Temporal Lobe Epilepsy. Neurosurgery. Dec 2016;79 Suppl 1:S83–S91. doi:10.1227/NEU.0000000000001439

27. Donos C, Rollo P, Tombridge K, Johnson JA, Tandon N. Visual field deficits following laser ablation of the hippocampus. Neurology. Mar 24 2020;94(12):e1303–e1313. doi:10.1212/WNL.0000000000008940

28. Salehi A, Yang PH, Smyth MD. Single-center cost comparison analysis of stereoelectroencephalography with subdural grid and strip implantation. J Neurosurg Pediatr. Feb 18 2022:1–7. doi:10.3171/2022.1.PEDS21523

