## Supplementary figures and images for "Laser Ablation of Periventricular Nodular Heterotopia for Medically Refractory Epilepsy"

### Supplemental Figure 1

a. imaging characteristics-LITT population

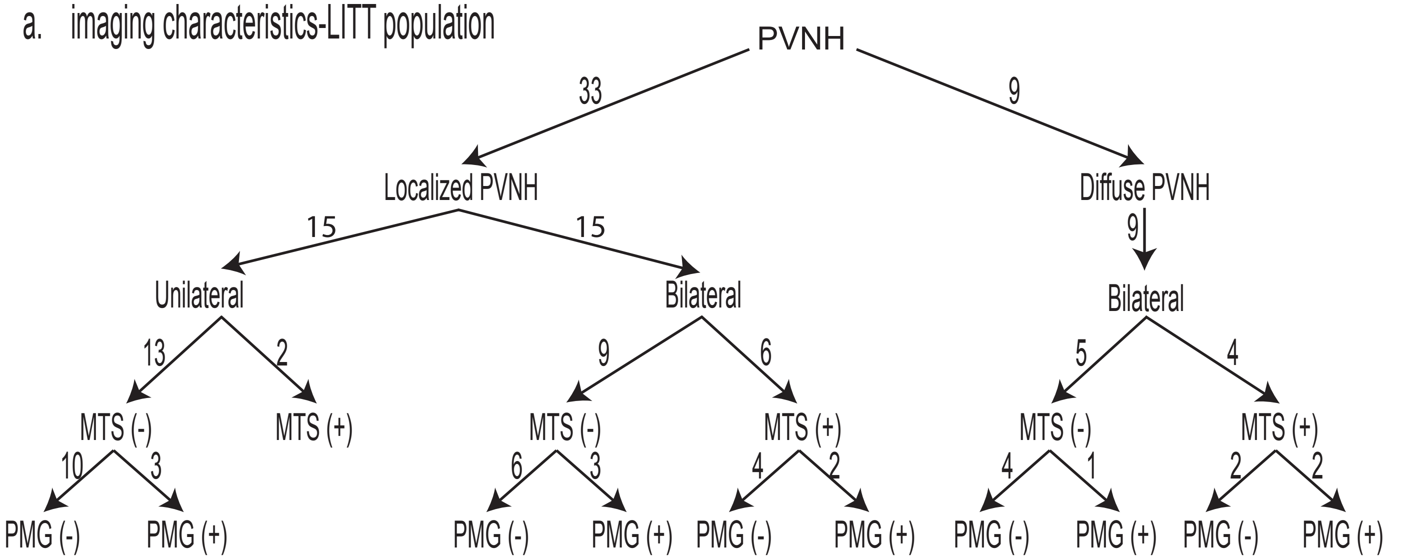
