## Supplemental table for "Laser Ablation of Periventricular Nodular Heterotopia for Medically Refractory Epilepsy"

Supplemental Table 1. Epilepsy characteristics and non-invasive workup of patients

| Pt #/ Gen | ASMs | | Seizure Semiology | Scalp EEG-Interictal | Scalp EEG-Ictal onset | | Imaging Findings | |
| --- | --- | --- | --- | --- | --- | --- | --- | --- |
| 1/M | 3 | FIAS | | R anterior temporal | | R anterior temporal | | B localized temporal occipital PVNH, R occipital PMG, R MTS |
| 2/M | 3 | FIAS | | R temporal | | R temporal | | B diffuse PVNH R>L; L PMG; B MTS |
| 3/F | 2 | FIAS, visual aura | | B anterior temporal | | R anterior temporal | | R localized occipital, temporal PVNH, R PMG, R intraparietal sulcus, closed-lip schizencephaly |
| 4/F | 2 | FTBTC | | L occipital | | L occipital | | B localized temporal, occipital; L PMG |
| 5/F | 2 | FTBTC | | B temporal | | R temporal | | R localized occipital, temporal PVNH |
| 6/M | 5 | FTBTC | | B frontotemporal | | R frontotemporal | | L localized temporal PVNH, schizencephaly |
| 7/F | 6 | FTBTC | | R frontal and temporal; L frontal and occipital | | L frontal | | B localized parietal, occipital, frontal, temporal |
| 8/M | 1 | FTBTC | | B temporal | | B temporal | | B Localized temporal, occipital, parietal (R>L) PVNH, L MTS |
| 9/F | 5 | FTBTC | | Normal | | No EEG correlation | | L Localized Frontal PVNH |
| 10/M | 2 | FIAS | | B temporal (L>R) | | L temporal | | B Diffuse PVNH; B MTS; R PMG |
| 11/M | 3 | FTBTC | | L temporal | | L temporal | | L localized temporal; B MTS |
| 12/F | 5 | FTBTC | | L frontotemporal | | L occipital | | L localized occipital PVNH |
| 13/F | 8 | FTBTC | | Normal | | B frontal, parietal | | B localized occipital PVNH, B MTS |
| 14/M | 3 | FIAS | | L temporal | | L temporal, R temporal | | R localized occipital PVNH, L MTS |
| 15/F | 6 | FIAS | | L temporal | | L temporal | | L localized occipital, temporal PVNH; L PMG |
| 16/F | 4 | FIAS; Rare FTBTC | | B temporal | | L temporal | | B localized frontal, parietal PVNH |
| 17/F | 4 | FTBTC | | L temporal | | L temporal | | B Diffuse PVNH (L>R) |
| 18/M | 8 | FTBTC | | R temporal; R parieto-occipital | | R parieto-occipital; R temporal | | R parieto-occipital FCD, R MTS, R localized occipital, temporal PVNH |
| 19/F | 2 | Visual aura | | L temporal | | No EEG correlation | | L localized occipital PVNH |
| 20/M | 3 | FTBTC | | B frontotemporal | | R temporal | | B diffuse PVNH |
| 21/F | 4 | FTBTC | | B frontal and temporal | | L frontal, right temporal | | B diffuse PVNH |
| 22/F | 1 | FIAS | | L temporal | | L temporal | | L localized temporal PVNH |
| 23/F | 3 | FIAS; Rare FTBTC | | B frontotemporal | | L frontal | | B localized temporal occipital PVNH |
| 24/F | 3 | FIAS; Rare FTBTC | | L temporal | | L temporal | | B localized frontal, parietal; L PMG |
| 25/F | 3 | FIAS | | L temporal | | Left temporal | | L localized parietal, temporal, occipital PVNH |
| 26/F | 3 | Visual aura | | Right temporal | | No EEG correlation | | R localized temporal occipital PVNH |
| 27/F | 5 | FTBTC | | B temporal | | R frontotemporal | | B diffuse PVNH; R MTS |
| 28/F | 6 | FTBTC | | L frontal and temporal | | R frontal | | B localized temporal, occipital, parietal; R PMG |
| 29/M | 2 | FIAS; Rare FTBTC | | L frontal and temporal | | L temporal; R frontal | | B localized temporo-occipital; B temporal PMG (R>L) |
| 30/F | 3 | FTBTC | | B temporal | | Left temporal | | B diffuse PVNH, R PMG, R schizenchepaly |
| 31/F | 4 | FTBTC | | L frontal R temporal | | R temporal | | R localized frontal PVNH; L MTS |
| 32/F | 5 | FTBTC | | B frontotemporal | | R temporal | | B localized occipital, parietal, temporal PVNH, B MTS |
| 33/M | 3 | FTBTC | | L temporal | | No EEG correlation | | B localized frontal, parietal, temporal; B MTS; L PMG |
| 34/M | 3 | FTBTC | | L temporo-parietal, R temporal | | R temporal | | B localized occipital, temporal PVNH; L PMG |
| 35/F | 6 | FTBTC | | B temporal (L>R) | | L temporal | | B diffuse PVNH; B MTS |
| 36/M | 5 | FIAS | | B frontotemporal | | L temporal | | B localized parietal, temporal PVNH (L>R) |
| 37/F | 3 | FTBTC | | B frontotemporal | | L temporal | | B diffuse PVNH |
| 38/M | 2 | FTBTC | | L frontotemporal | | L temporal | | L localized temporal, occipital PVNH |
| 39/F | 1 | FTBTC | | B temporo-parietal | | L temporal | | B localized occipital, parietal, temporal PVNH |
| 40/F | 6 | FIAS | | R temporal, R parietal | | R temporal | | R localized frontal, parietal, occipital PVNH |
| 41/F | 2 | FIAS; Rare FTBTC | | R frontal | | R frontal | | R localized temporal, occipital, parietal PVNH |
| 42/F | 2 | FIAS | | R temporal | | R temporal | | R localized temporal, atrial PVNH; R parietal PMG |

ASM: anti-seizure medications; FIAS: Focal impaired awareness seizure; FTBTC: Focal to bilateral tonic clonic seizure; sEEG: stereo-electroencephalography; SDE: Subdural electrodes; F: Female; M: Male; L: Left; R: Right;

Supplemental Table 2: Invasive evaluation, ablation percentage and seizure outcomes

| Patient | Intracranial evaluation | SEEG interictal | SEEG ictal | LITT Side  /# Fibers | | % Ablation PVNH (SoZ) | Last Engel score/ILAE score Months Postop | | | # AED at last FU |
| --- | --- | --- | --- | --- | --- | --- | --- | --- | --- | --- |
| 1 | B sEEG | R PVNH, R AH, L PVNH | L PVNH and cortex; R AH; R PVNH and cortex | R | 1 | 97.5 | Id | 1 | 85 | 1 |
| 2 | B sEEG | L PVNH; R AH | L PVNH; PVNH to hippocampus | L | 2 | 99.2 | Ia | 1 | 77 | 1 |
| 3 | R sEEG | R AH, R PVNH | R PVNH and overlying cortex -> spread | R | 2 | 99.7 | Ia | 1 | 72 | 0 |
| 4 | B sEEG followed by L SDE | L PVNH | L PVNH | L | 2 | 93.2 | Ia | 1 | 70 | 1 |
| 5 | R sEEG | R AH; R PVNH | R PVNH; R PMG | R | 2 | 100 | Ia | 1 | 67 | 0 |
| 6 | B sEEG | R SMA; R insula; L PVNH | R SMA/Insula; L motor cortex | -- | -- | n/a—R RNS | IVb | 4 | 64 |  |
| 7 | B sEEG followed by R SDE | B PVNH | L PVNH | B | 4 | 95 | Ia | 1 | 63 | 4 |
| 8 | B sEEG | R PVNH; R Hip; L Hip | L hippocampus; R PVNH -> R AH | R | 1 | 98.9 | Ia | 1 | 56 | 0 |
| 9 | L sEEG and L SDE | L PVNH | L PVNH | L | 1 | 100 | Ic | 1 | 48 | 2 |
| 10 | B sEEG and L SDE | L PVNH; L AH | L posterior temp PVNH with spread to mesial temporal; L hippocampus to mesial temporal | L | 2 | 94 | IIc | 3 | 44 | 2 |
| 11 | L sEEG | L AH, L temp; L ant PVNH | L Ant PVNH -> AH | L | 1 | 97.6 | Ic | 1 | 39 | 2 |
| 12 | B sEEG | L temp PVNH; L AH | PVNH to temporal/insula | L | 2 | 95.1 | IIb | 3 | 37 | 2 |
| 13 | B sEEG followed by R SDE | L AH-> opercular; R PVNH | R occipital PVNH; R temporal PVNH | B | 2 | 100 | Ia | 1 | 26 | 3 |
| 14 | B sEEG | R PVNH; L AH | L AH, R AH, L insula, L orbitofrontal | -- | -- | n/a—VNS | IVa | 4 | 54 |  |
| 15 | L sEEG | L atrial PVNH | L AH; L atrial PVNH | L | 1 | 95.1 | IIb | 3 | 12 | 2 |
| 16 | B sEEG | L PVNH | L atrial PVNH | L | 1 | 82.9 | IIIa | 4 | 25 | 2 |
| 17 | B sEEG | L AH; L temp | L AH; L atrial PVNH | L | 2 | 94.1 | Ia | 1 | 27 | 3 |
| 18 | R sEEG | Occipital FCD | Occipital FCD | -- | -- | n/a—occipital topectomy | IVb | 4 | 50 |  |
| 19 | L sEEG | L PVNH | L PVNH | L | 1 | 97.3 | IIc | 3 | 36 | 1 |
| 20 | B sEEG | R occipital/Atrial PVNH; L temporal PVNH | L temporal PVNH | B | 4 | 82.5 | IIIa | 4 | 37 | 1 |
| 21 | B sEEG | L atrial PVNH, R ant temporal PVNH -> temp pole; | L PVNH; R temp PVNH | B | 4 | 83 | IVb | 5 | 26 | 4 |
| 22 | L sEEG | L AH | L PNVH -> AH | L | 1 | 100 | Id | 1 | 26 | 1 |
| 23 | B sEEG | L PVNH -> cortical spread | L PVNH | L | 1 | 78.8 | IIIa | 4 | 12 | 2 |
| 24 | B sEEG | L frontal -> PVNH; L AH | L PVNH -> L cortical spread | L | 1 | 95.6 | Ia | 1 | 26 | 0 |
| 25 | B sEEG | L AH; L PVNH | L PVNH -> AH | L | 1 | 96.8 | Ia | 1 | 25 | 2 |
| 26 | R sEEG | R temporal PVNH | R temp PVNH with cortical spread | R | 1 | 99.3 | Ia | 1 | 18 | 0 |
| 27 | B sEEG | R temp PVNH; R AH | R temporal PVNH | R | 1 | 100 | Ib | 2 | 13 | 1 |
| 28 | B sEEG | R atrial, occipital PVNH, R PMG | R PVNH | R | 1 | 99.2 | Ib | 2 | 24 | 2 |
| 29 | B sEEG | L occipital PVNH; R Atrial PVNH, PMG | R atrial PVNH; R frontal PVNH; L occipital -> Hippo; PMG | B | 4 | 92 | Ia | 1 | 24 | 2 |
| 30 | B sEEG | L AH/insula; L PVNH | R atrial PVNH; L PVNH | B | 4 | 91 | IIb | 3 | 24 | 3 |
| 31 | B sEEG | R atrial PVNH | R PVNH -> AH | R | 2 | 89.4 | Ib | 2 | 24 | 1 |
| 32 | B sEEG | R atrial PVNH | R atrial PVNH;  L temporal PVNH -> Hippo | B | 2 | 94.8 | Ia | 1 | 24 | 0 |
| 33 | B sEEG | L PVNH; L PMG; L AH | L PVNH -> AH | L | 2 | 96 | Ib | 2 | 15 | 1 |
| 34 | B sEEG | L PVNH; L pmg | L PVNH -> AH;  L PMG; R atrial PVNH | B | 3 | 100 | Id | 1 | 15 | 2 |
| 35 | B sEEG | L AH; R atrial PVNH | R atrial PVNH; L temporal PVNH -> AH/temporal pole | B | 2 | 98 | Ia | 1 | 13 | 2 |
| 36 | B sEEG | L atrial PVNH | L atrial PVNH | L | 1 | 91.1 | Id | 3 | 15 | 2 |
| 37 | B sEEG | R frontal PVNH; L parietal PVNH | R PVNH | B | 3 | 89.9 | Ia | 1 | 14 | 2 |
| 38 | L sEEG | L occipital PVNH | L occipital PVNH | L | 1 | 93.1 | Ia | 1 | 9 | 1 |
| 39 | B sEEG | L occipital PVNH | L PVNH;  R PVNH | B | 3 | 88.9 | Id | 1 | 25 | 1 |
| 40 | R sEEG | R parietal PVNH | R PVNH -> cortical spread | R | 2 | 96.6 | Ia | 1 | 12 | 0 |
| 41 | R sEEG | R frontal PVNH | R PVNH | R | 1 | 97.4 | Ia | 1 | 12 | 2 |
| 42 | R sEEG | R PMG; R atrial PVNH | R atrial PVNH, R PMG | R | 3 | 91 | Ia | 1 | 8 | 0 |

**Supplemental Table 3 Epilepsy Surgery Procedures other than LITT in this cohort**

| Patient | Procedure | SEEG vs. epilepsy surgery timing | Surgery vs. PVNH-LITT timing | Interval (Months) | Engel Class Prior to Additional Epilepsy Procedure | Engel Class Post  Additional Epilepsy Procedure |
| --- | --- | --- | --- | --- | --- | --- |
| 11 | LITT L SAH | Post | Pre | 36 | IVb | Ic |
| 15 | VNS | Post | Pre | 63 | IIIa | IIb |
| 19 | L ATL | Post | Pre | 58 | IVb | Iic |
| 27 | VNS | Post | Pre | 27 | IIIa | Ib |
| 33 | L ATL | Post | Pre | 36 | IIIa | Ib |
| 36 | VNS | Post | Pre | 84 | IVb | Id |
| 1 | R ATL with partial PVNH removal | Pre | Pre | 2 | Ia | Id |
| 2 | R ATL | Pre | Pre | 12 | IVb | Ic |
| 4 | L topectomy | Pre | Pre | 2 | IIIa | Ic |
| 7 | R topectomy | Pre | Pre | 3 | IIIa | Ia |
| 8 | LITT L SAH | Pre | Pre | 8 | IIIa | Ia |
| 17 | LITT L SAH | Pre | Pre | 6 | IVb | Ia |
| 24 | LITT L PMG | Pre | Pre | 18 | IVb | IIIa |
| 31 | R ATL | Pre | Pre | 12 | IVb | Ib |
| 10 | L ATL | Pre | Post | 13 | IIa | Iic |
| 28 | R topectomy | Pre | Post | 3 | Ia | Ia |
| 6 | RNS |  | No LITT | -- |  | IVb |
| 14 | VNS |  | No LITT | -- |  | IVb |
| 18 | Occipital Topectomy |  | No LITT | -- |  | IVb |

ATL: Anterior temporal lobectomy; VNS: vagal nerve stimulator; RNS: Responsive Neurostimulation; SAH selective amygalo-hippocampectomy

**Supplementary Table 4: Extent of SoZ ablation by LITT in patients with unilateral and bilateral onsets**

|  | Ablated % (Entire SoZ) | PVNH % (Near SoZ) | Cortical % (with known abnormality near SoZ) |
| --- | --- | --- | --- |
| **Unilateral SoZ** |  |  |  |
| - Nodule | N = 15 \| 97.7% | 97.7 | -- |
| - Nodule + Cortex (total) | N = 13 \| 94.5% | 94.5% | 93.1% |
| - N+C (good outcome) | N = 11 \| 94.7% | 96.2% | 93.1% |
| - N+C (poor outcome)* | N = 2 \| 80.8%* | 83.8% | -- |
| **Bilateral SoZ** |  |  |  |
| - Nodule | N = 6 \| 94.8 | 94.8 | -- |
| - Nodule + Cortex (total) | N = 5 \| 89 | 89.5 | 88.3 |
| - N+C (good outcome) | N = 3 \| 91.3 | 94.3 | 88.3 |
| - N+C (poor outcome)* | N = 2 \| 82.5 | 82.5 | -- |

* No lesional cortical abnormality identified; N: Nodule; C: Cortical

**Supplemental Table 5: Extent of total abnormal imaging ablation with unilateral and bilateral onsets**

|  | Ablated % (total abnormality) | PVNH % | Cortical % |
| --- | --- | --- | --- |
| **Unilateral Total Ablation** |  |  |  |
| Nodule | N = 15 \| 79.1% | 79.1% | -- |
| Nodule + Cortical (total) | N = 13 \| 69.5% | 73.8% | 63% |
| N+C (good outcome) | N = 11 \| 70.3 | 74.2% | 63% |
| N+C (poor outcome/  Engel 3/4)* | N = 2 \| 70.6* | 70.6 | -- |
| **Bilateral Total Ablation** |  |  |  |
| Nodule | N = 6 \| 71.2% | 71.2% | -- |
| Nodule + Cortical | N = 5 \| 53.9 | 60.3 | 49.3 |
| N+C (good outcome) | N = 3 \| 53.2 | 60.5 | 49.3 |
| N+C (poor outcome/  Engel 3/4)* | N = 2 \| 59.5* | 59.9 | -- |

* No lesional cortical abnormality identified; N: Nodule; C: Cortical
